# From 5Vs to 6Cs: Operationalizing Epidemic Data Management with COVID-19 Surveillance

**DOI:** 10.1101/2020.10.27.20220830

**Authors:** Akhil Sai Peddireddy, Dawen Xie, Pramod Patil, Mandy L. Wilson, Dustin Machi, Srinivasan Venkatramanan, Brian Klahn, Przemyslaw Porebski, Parantapa Bhattacharya, Shirish Dumbre, Erin Raymond, Madhav Marathe

## Abstract

The COVID-19 pandemic brought to the forefront an unprecedented need for experts, as well as citizens, to visualize spatio-temporal disease surveillance data. Web application *dashboards* were quickly developed to fill this gap, including those built by JHU, WHO, and CDC, but all of these dashboards supported a particular niche view of the pandemic (ie, current status or specific regions). In this paper^1^, we describe our work developing our own COVID-19 Surveillance Dashboard, available at https://nssac.bii.virginia.edu/covid-19/dashboard/, which offers a universal view of the pandemic while also allowing users to focus on the details that interest them. From the beginning, our goal was to provide a simple visual way to compare, organize, and track near-real-time surveillance data as the pandemic progresses. Our dashboard includes a number of advanced features for zooming, filtering, categorizing and visualizing multiple time series on a single canvas. In developing this dashboard, we have also identified 6 key metrics we call *the 6Cs standard* which we propose as a standard for the design and evaluation of real-time epidemic science dashboards. Our dashboard was one of the first released to the public, and remains one of the most visited and highly used. Our group uses it to support federal, state and local public health authorities, and it is used by people worldwide to track the pandemic evolution, build their own dashboards, and support their organizations as they plan their responses to the pandemic. We illustrate the utility of our dashboard by describing how it can be used to support data story-telling – an important emerging area in data science.

## I. Introduction

The COVID-19 outbreak caused by the novel coronavirus SARS-CoV-2 has disrupted the lives of people globally. It has had a huge impact on health, economies, and society in general, undoubtedly making it the pandemic of the century. As of October 25, 2020, the cumulative number of confirmed COVID-19 cases exceeded 43 million worldwide, with almost 1.15 million deaths. With the uncertainty surrounding the pandemic and its impact, there has been a great need for surveillance of pandemic data in order to make informed and fact-based decisions. The tracking of the pandemic data has been of prime importance for policymakers, public health officials, and academic researchers attempting to understand and respond to this public health crisis, along with every layperson concerned about how the pandemic will affect their daily life. Many dashboards have been developed to help the public better understand the current status, each focused on a different aspect of the pandemic. However, there is no gold standard set for epidemic data management and visualization. There is a very clear need for a user-centric, one-stop solution backed by a reliable source of data and coupled with rich visualizations and an easy-to-use interface that is accessible to both the public and researchers.

These issues led us to identify 6 metrics that might serve to define a standard for epidemic surveillance data management that we call *the 6Cs standard*. The 6Cs standard proposes that epidemic surveillance data should be *Consistent, Correct, Current, Comprehensive, Curated, and Computer-readable*. With this standard in mind, we created a COVID-19 surveillance dashboard which offers data exploration and visualization features designed to assist researchers, but which even a normal user, unfamiliar with the technical details, can understand. While we sincerely appreciate the contributions and efforts put forth by teams working on similar dashboards, we have observed that these sources are either limited in scope or miss some of the characteristics of the 6Cs standard. Our goal is to provide an insightful view into COVID-19 incidence data through spatio-temporal surveillance visualizations conforming to the 6Cs standard.

The Biocomplexity Institute & Initiative’s COVID-19 Surveillance Dashboard, which was released on February 3, 2020, is available at https://nssac.bii.virginia.edu/covid-19/dashboard/. It is a single-page, interactive and responsive web application that is dynamically updated; it allows end users to view and explore COVID-19 case counts at both temporal and spatial resolutions with just a few mouse clicks. To the best of our knowledge, our dashboard is one of very few that presents historical data in three visualization formats: choropleth map (Fig. 1a), charts (Fig. 5a), and a data table, all of which are interactive. Our charts include Cumulative and Incident Epicurves for all regions, and our unique use of a movie-style time slider makes it easy to simultaneously explore both temporal and spatial evolution of the pandemic. In its current form, the dashboard supports data rendering at the country level for all countries in the world; at the state and province level for 20 countries; and county-level statistics for the United States of America (USA). This data can be easily accessed either through the query tool, by filtering the data, or by clicking on the zoomable map. As of October 25, 2020, more than 1.13 million users from over 220 countries have used our dashboard, and more than 60 million requests were processed on the main feature layer hosted on ArcGIS Online.

**Fig. 1:**
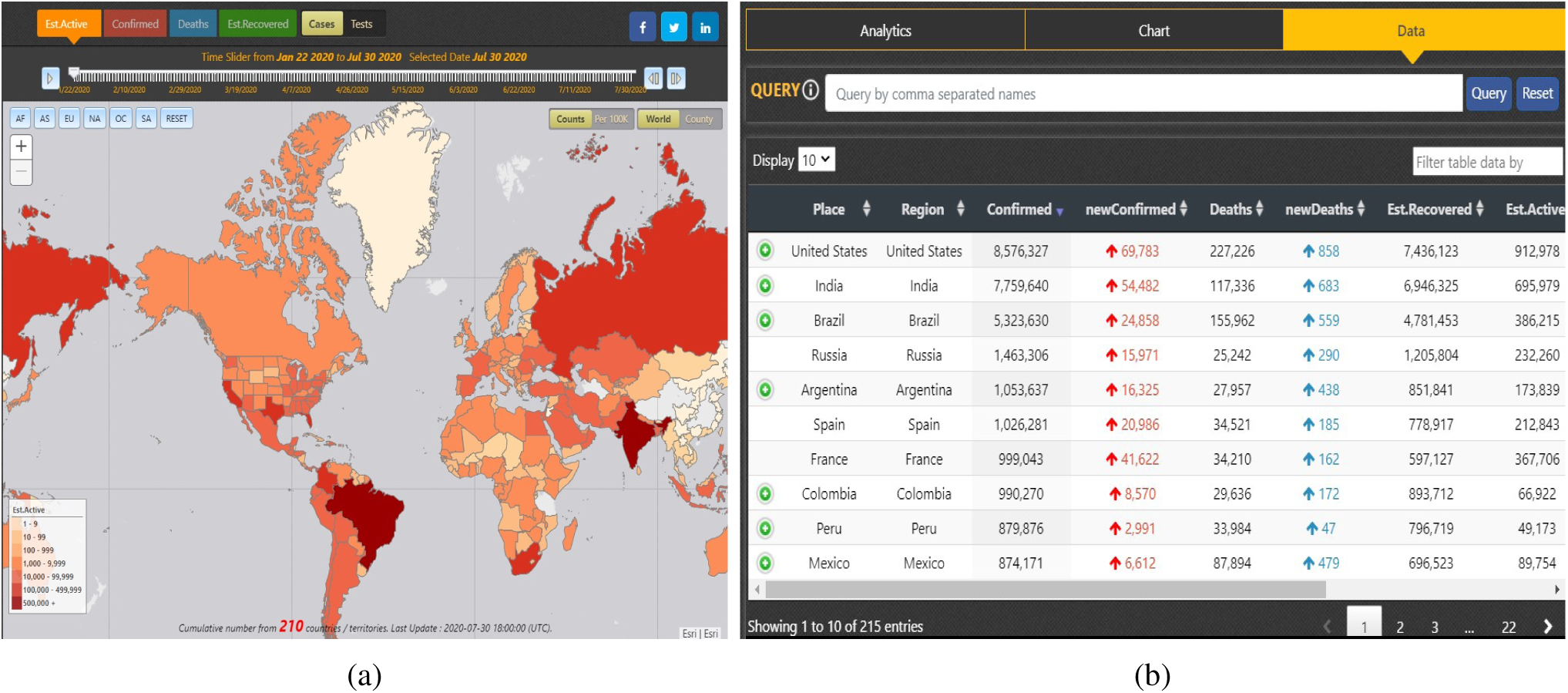
Dashboard screenshots. (a) Map panel. Choropleth map of the world, rendered with the estimated active count, but with the option of switching to other layers: state/province layer, county layer, or different attributes for rendering. (b) Information panel - Data Table. This includes interactive data table with data of different attributes like Region, Confirmed, Deaths, Est.Recovered, Last Update etc.

**Fig. 2:**
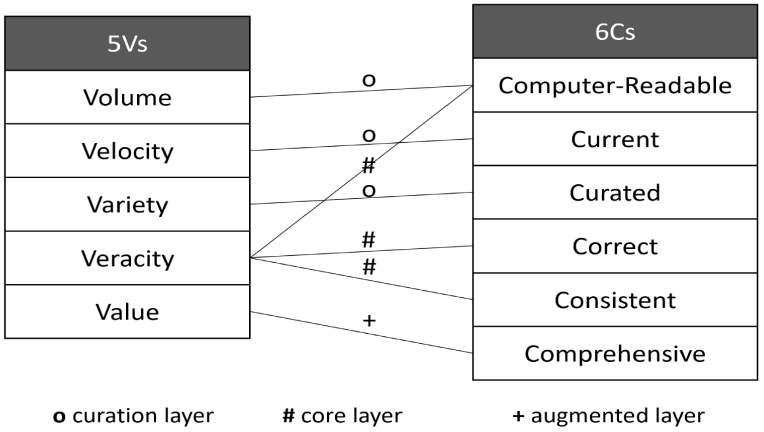
Relationship between the 6Cs and the 5Vs of Big Data, and the data pipeline layers used to achieve them in our surveillance dashboard.

**Fig. 3:**
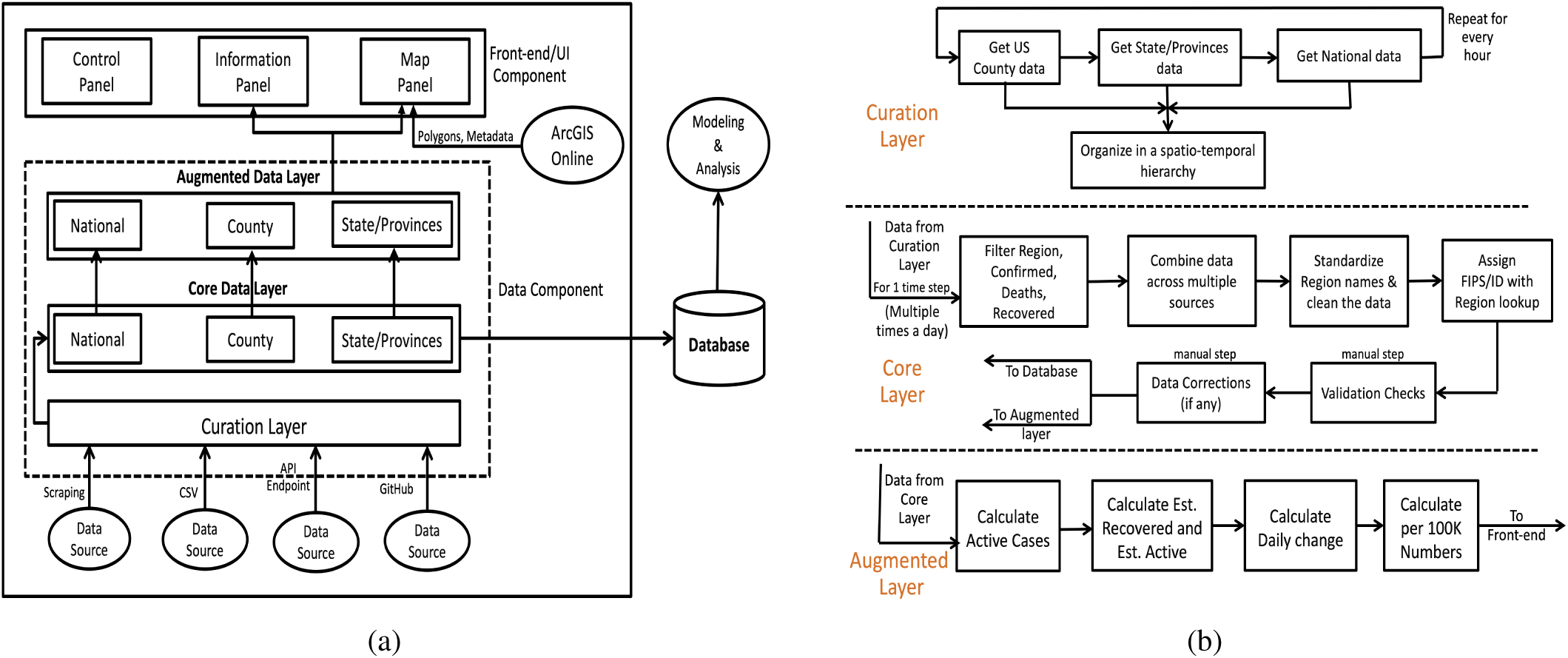
(a) Dashboard architecture: The UI consists of a control panel, information panel and map panel. The data component is a pipeline with three layers: curation layer, core data layer and augmented data layer. (b) Micro services used in the surveillance data pipeline. The series of steps that take place in the data component’s layers, from the collection of data to its organization, integration, validation and augmentation in order to facilitate its usage in modeling and the front-end UI.

**Fig. 4:**
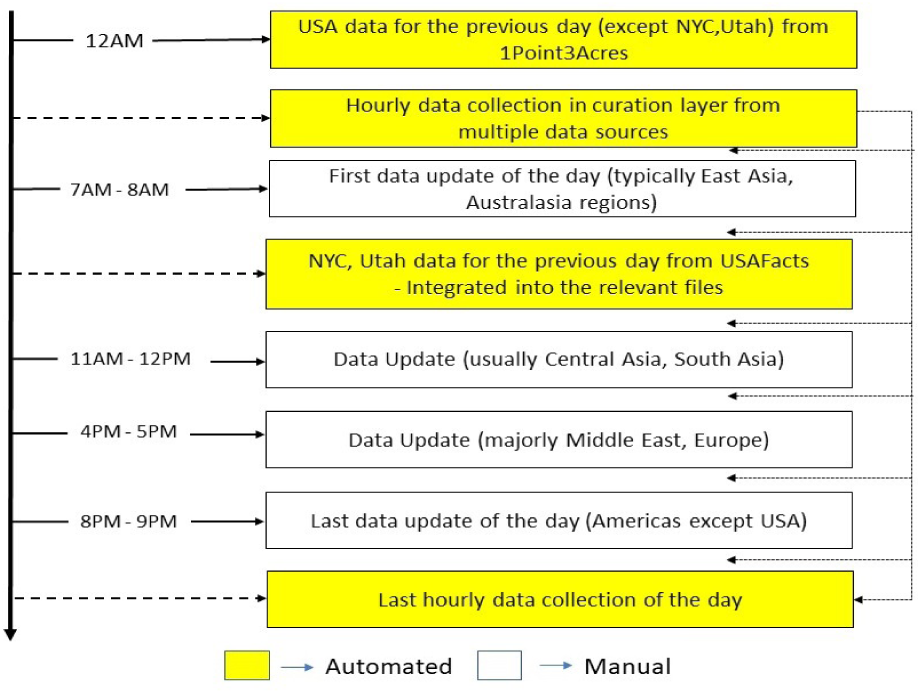
Timeline of daily data updates. This sequence illustrates the 24-hour data update workflow, which is a combination of automated & manual steps. It includes details on which regions’ data typically gets updated in each round.

**Fig. 5:**
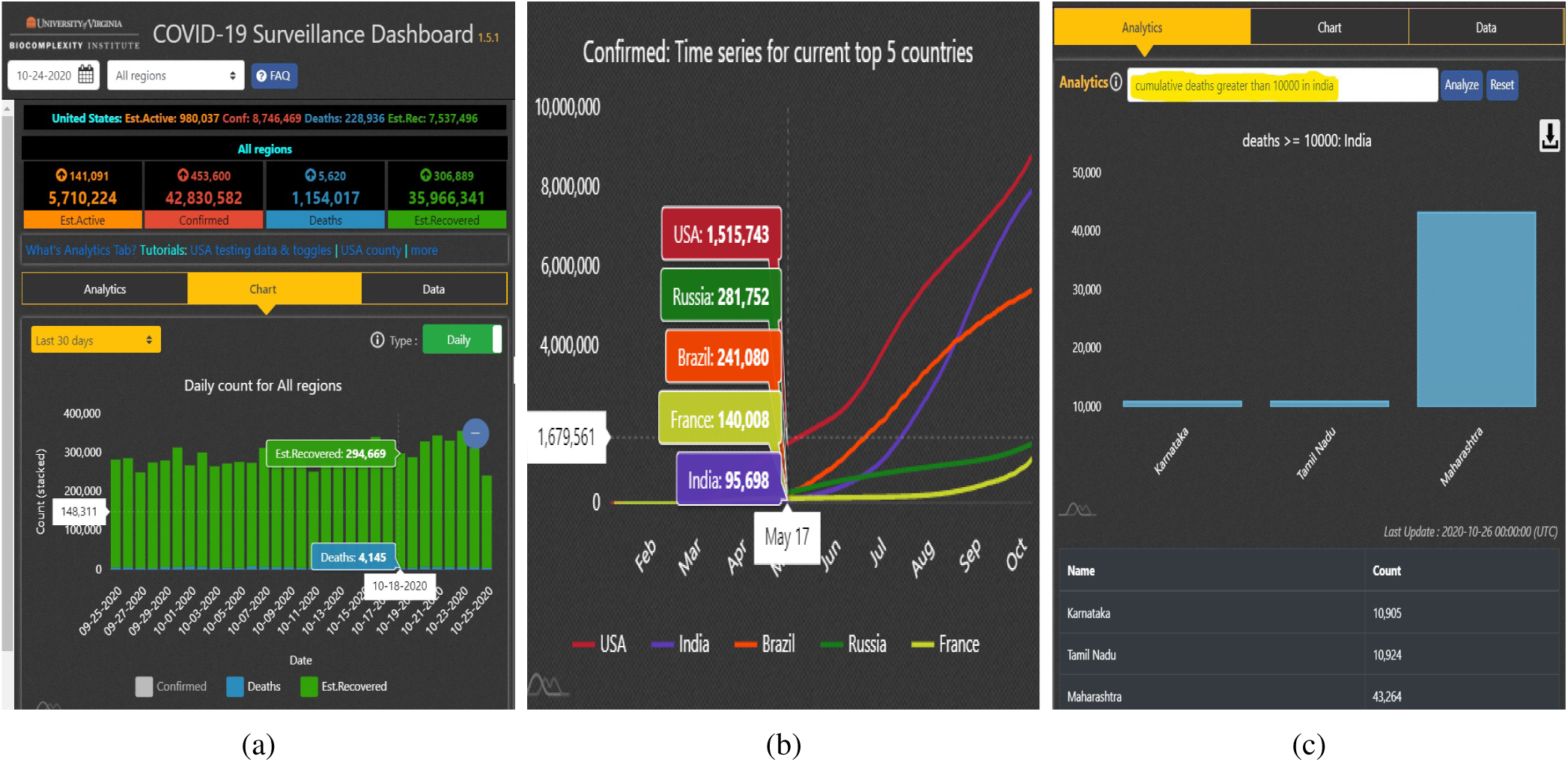
User Interface. (a) Information panel - Charts. This shows interactive charts for cumulative & incidence data for different attributes and summary statistics. (b) Top K charts. The plot to compare the epicurves of the top k regions for different attributes, either by cumulative or incidence data. (c) Information Panel - Analytics. Analytics through Question Answering. Includes results with data and chart.

In this paper, we discuss the importance of the 6Cs of epidemic data management and why it should be treated as a standard, followed by a discussion on its relationship with the 5Vs of big data. We also provide an overview of other COVID-19 and epidemic dashboards in the related work section. We then describe the procurement and curation of the dashboard data, including the sources used and our algorithm for estimation of COVID-19 recoveries - a very unique feature of our dashboard. We discuss the underlying architecture of our dashboard, along with user interface (UI) implementation details. Finally, we discuss the utilities of our dashboard, the challenges faced, and lessons learned.

## II. The Importance of the 6Cs Standard

### A. Consistent

Consistency can be viewed from two perspectives - consistency in the format of the data, and consistency in the content of historical data. One of the major goals of collecting and managing epidemic data is to support informed health and public policy decisions. This is applicable in various contexts, including policymakers and the academic researchers who support them, individual businesses, and the general public. This decision-making process often depends on projections or forecasts of the pandemic spread produced by epidemic modeling simulations, which, in turn, depend on surveillance data. Modeling applications expect that the data format will be consistent over time; frequent format changes would necessitate frequent revisions to model implementations, causing untimely delays in forecast production. This uncertainty may lead modelers to seek more consistent data sources. Policymakers and public health officials also depend on websites and dashboards to guide their decisions. Many of these online tools, including ours, depend on open data sources. If the underlying format of these sources changes, it can delay dashboard updates, which may, in turn, prevent the policymakers from having the most current information when they need it.

Another perspective is consistency of the historical data. Once published, historical data should be updated as little as possible, except in the case when a previous entry is discovered to be invalid. To reduce the risk of data contamination, data should be collected from proven reliable sources; collation and correction of the data needs to be performed via a consistent and predictable process; and frequent validation must be a critical part of the curation process. In the event that a correction is made, the downstream impact on forecasts and projections could be high, so it is necessary to provide a record of data updates that is accessible to all consumers of the data. This also leads to our next C, which is Correctness.

### B. Correct

Epidemic data is sensitive and inaccuracies can have a catastrophic impact on public health. This raises the importance of the data correctness. Large scale data curation is prone to error that occurs for a variety of reasons, including incorrect data entry, improper access to the data, or errors in processes performing calculations and data wrangling. Steps must be taken to reduce data error to the maximum extent possible. This can be achieved through proper validation and data checks; for example, the cumulative case counts should not decrease unless there have been upstream revisions of historical data by the original reporting source, or the total count of a region should be equal to the summation of its subregions, etc. Any uncertainty that cannot be resolved should be explicitly accounted for and documented.

### C. Current

A pandemic such as COVID-19 evolves quickly. We have all seen situations where a region with no prevalence of infections suddenly emerges as a hotspot with a quick case doubling time. Hence, the frequency of data updates is vital, as stale data does not capture the current status, and is a poor guide for making decisions under rapidly changing conditions. This emphasizes the need for timely updates, which, in turn, requires automated data collection and maintenance of historical snapshots of the data. It also amplifies the need for storing the data in a temporal representation with a clear indication of when data updates have occurred.

### D. Comprehensive

A dashboard tracking an epidemic should be comprehensive in providing detailed visual analysis with charts, geospatial mapping, and time series visualizations, as well as with summary statistics. Apart from that, several other metrics can be derived from the core data of cases and deaths to help users understand the present situation in a clear manner. For instance, active cases is an important metric in assessing the current status of the disease, and the number of infections normalized by the population demonstrates the density of the spread. Other data, like laboratory testing, hospitalizations, mobility, and interventions, have a direct influence on the pandemic and add meaning to the core data. Having a comprehensive, complete picture of the pandemic can help researchers understand which factors could curtail the spread of the disease.

### E. Curated

The epidemic data should be curated from diverse sources in order to cater to the large and disparate needs of the population. First, the data should be available for as many regions and subregions as possible to get a more global picture of the disease spread. This not only makes the dashboard complete and serves the majority of the users, but also helps improve decision-making at the local level. The kind of interventions required for a nation with multiple hotspots and a nation with a single adversely affected hotspot are quite different, which can only be captured if data is available at multiple levels of spatial resolution.

### F. Computer-readable

To cater to downstream tasks like modeling, analysis or visualization, data should be Computer-readable, meaning it should be easily accessible in the form of CSV files, databases, or through an API endpoint. Data provided in a textual format, such as a report or an article, is good for human consumption, but requires a lot of preprocessing and manual work to prepare it for other computational tasks. Similarly, data should also have standard geospatial mapping and naming conventions in order to easily identify or differentiate between the regions and subregions. In addition, hierarchical organization of the spatial and temporal components in the data also facilitates data retrieval.

### G. The 5Vs of Big Data

Volume, Velocity, Variety, Veracity, and Value are popularly known as the 5Vs of Big Data. Although the 6Cs of epidemic data management bear some resemblance to the 5Vs, the two standards actually complement each other quite a bit (See Fig. 2)

Although the size of epidemic data, specifically for an emerging infectious disease, is not as large as that in a typical “Big Data” setting, epidemic data still has a lot of spatial and temporal components. As a pandemic progresses, the size of the data increases rapidly, and when additional data types like mobility, tests, and hospitalizations are added to the set, scalability becomes an important factor. Computer-readability plays an important role in promoting efficient handling of the high Volume of multidimensional data, while still supporting flexibility and easy accessibility. The characteristic Velocity resembles Current, specifically with the temporal component where the data flows continuously from multiple sources into the application for real-time updates. Several optimization techniques, along with a minimal amount of human intervention, can help to handle the Velocity of data. Epidemic data has a wide Variety of potential data sources and formats, ranging from a structured form like CSV, to a semi-structured form like Rest API endpoints and dashboards, to, finally, unstructured forms like webpages or reports. This is ideally what Curated in the 6Cs aims to handle, specifically for the spatial component. Veracity refers to reconciliation of inconsistencies and uncertainty in the data. Collected epidemic data is typically unstructured and messy, so it has to be cleaned and validated to attain Consistency and Correctness, with the right amount of data – not more and not less – because the data has no Value by itself unless it is refined down to include only the useful information. The Comprehensive characteristic helps useful information to be conveyed through rich visualizations and analysis.

## III. Related Work

In this section, we present an overview of some well-known COVID-19 dashboards and efforts from various groups for epidemic data management and visualization.

When we started our dashboard in late January, the Center for Systems Science and Engineering at Johns Hopkins University (JHU CSSE) [1] was one of the few organizations that gathered and shared COVID-19 data through a dashboard. As critical and influential as their data collection and curation efforts were, especially at the beginning of the pandemic, their data format and sharing platform has changed frequently, which has made it difficult for downstream users to adapt. This drove home the need for data Consistency as one of the most important goals to pursue. Another limitation of the JHU CSSE dashboard is the lack of temporal data and region-level visualizations, along with the inability to query or search for a specific region. The data for each category of cumulative data (cases, deaths, recovered etc.) is organized into separate tabs and panels, which makes it difficult to assess the full picture for a particular region.

1Point3Acres’ Global COVID-19 Tracker & Interactive Charts [2] dashboard initially focused on providing near real-time case information for North America. We were one of the early adopters of their data, and have used it for USA since early March. 1Point3Acres has added rich visualizations and expanded data coverage to more countries over time, but they still don’t cover all of the countries which have cases. Although 1Point3Acres provides spatial rendering for USA and Canada, this component is missing for other countries. The temporal data and visualization is also unavailable at sub-national levels, i.e. for states, provinces, and counties. Because of these shortcomings, this dashboard does not meet the 6Cs standards for Comprehensive and Curated.

Worldometer’s COVID-19 website [3] collects national-level data from every country and makes it available in a data table for 3 days. For USA, they provide detailed case counts down to the county level. They also display charts of historical data, which allow users to understand the current status of the pandemic, as well as some aspects of how it evolved over time. However, their data presentation is largely text-based (with the exception of the historical charts), and does not provide spatial visualizations, like maps, that would allow users to visualize differences between contiguous regions. For these reasons, Worldometer falls short of our standard for Comprehensive.

Public health organizations like the World Health Organization (WHO), Centers for Disease Control and Prevention (CDC), and the European Centre for Disease Prevention and Control (ECDC) [4]–[6] provide data that is Consistent, Correct, Current, and Computer-readable, but not completely Curated because they support data at only a single spatial resolution. WHO and ECDC provide the data for all the countries of the world, but not for states, provinces, or counties, whereas CDC provides data for USA states and counties, but not for other countries and subregions.

A number of dashboards have been developed that are specific to a region or a certain area of study. For example, health departments of many countries, states and counties have their own dashboards to track the local pandemic situation. Wissel et al. [7] used an R Shiny app for surveillance of USA cities using data from JHU CSSE, The New York Times, and the COVID Tracking Project. Barone et al. [8] developed a statistical surveillance dashboard based on their analysis of the ratio between cases and the days since the first case in the countries to determine the average speed of its epidemic motion, analogous to concepts in physics. Hohl et al. [9] built an R Shiny app based on their study of space-time scan statistics to detect daily clusters at the county level. On the topic of data curation, there are several efforts that collate data types other than surveillance data, like BeOutbreakPrepared [10] which provides individual-level epidemiological data, also known as line lists.

In the development of our dashboard, the Single Page Application (SPA) and movie-style time slider for temporal visualization of the surveillance data was inspired by similar functionality that was available in the now-defunct EpiViewer application [11]. The division of the front end into several panels is also similar to the presentation provided by EpiViewer, but we extended that concept with the addition of a geographical component and near real-time data from the current outbreak.

## IV. Data and Sources

This section describes the data elements our dashboard depends upon. Although our full dataset is not currently downloadable from our dashboard, we do maintain a Consistent data structure across all regions. We don’t conduct our own surveillance activities, so we rely on reputable, openly available data sources to populate our dashboard; those sources are also described in detail in this section.

### A. Data Fields

#### Core Surveillance data

Sub-Region (if any), Region, Confirmed Cases, Deaths, Reported Recovered. (Cumulative numbers are provided for the three numeric measures).

#### COVID Testing Data

Sub-Region (if any), Region, Positive Tests, Negative Tests, Total Tests, Positivity Rate (%), Data Quality Grade.

#### Augmented Surveillance Data

- Active cases : Confirmed - Deaths - Reported Recovered;
- ID/FIPS : ISO3 for countries, FIPS for USA counties and ID for states / provinces (hereby referred to as admin1 regions) by mapping these regions with an ISO lookup;
- Coordinates : Latitude and Longitude for GIS is obtained from ID/FIPS;
- Last Update : The UTC time when the data was last fetched and updated;
- Estimated Recovered : Estimate of Recovered case count calculated based on the time series of confirmed cases and deaths (more on our algorithm below);
- Estimated Active : Confirmed Cases - Deaths - Estimated Recovered;
- New Cases, New Deaths, New Recovered, New Estimated Recovered, New Estimated Active : Increase in counts from previous day’s cumulative numbers (Incidence Data);
- Per 100K counts : Population-normalized numbers for all of the above relevant data fields.

#### Estimating Recovered Counts

This feature is unique to our dashboard. A significant number of countries or states do not report the number of people who have recovered from COVID-19, and those that do report these numbers are not always up-to-date. The United Kingdom (UK), Netherlands, and Sweden are some of the countries which don’t report recovered statistics, and this data is not available for the majority of the states, provinces and counties performing independent reporting. Without knowing the number of recoveries, it is very difficult to calculate the number of Active cases, which is arguably a more important metric to track than Confirmed cases; for example, many local governments use active case counts to plan their reopening strategies. Furthermore, inaccurate recovery counts will lead to inaccurate active case counts. This raises the need for a well-defined method for calculating the number of recovered cases, and, by extension, active cases, which is consistent across all regions. Such an approach will minimize differences in reporting, hence allowing for fair comparison across regions.

To this end, we developed an algorithm for calculating the number of recovered cases. A joint study conducted by WHO and China [12] concludes that the median time from onset of COVID-19 to clinical recovery for patients with mild cases is approximately 2 weeks, while the median time is 3 to 6 weeks for patients with more severe or critical disease symptoms. A cohort study by Wu Z, McGoogan JM [13] shows that 81% of cases are mild to moderate, 14% are severe, and 5% are critical. This study is referenced by the official CDC interim clinical guidance [14]. Illinois Department of Public Health follows a similar estimate for their calculation of recovered cases [15]. Based on these studies, we calculate Estimated Recovered as follows:

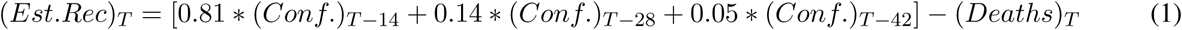

where T represents the day in the time series for which Estimated Recovered is calculated. While this is a fairly safe estimate for the number of recovered cases, it is possible that actual recovery counts will vary depending on the region or subregion. In cases where the reported recovery number is higher than our safe estimate, we set Estimated Recovered equal to the reported value. If the CDC or WHO guidelines regarding the recovery estimates are updated, we will adjust our formulas accordingly.

### B. Data Sources

#### Epidemic surveillance data

We use several data sources for collating our epidemic surveillance dataset. Table I summarizes which data sources we are currently using, how often we poll those sites, and the collection methods.

**TABLE I:**
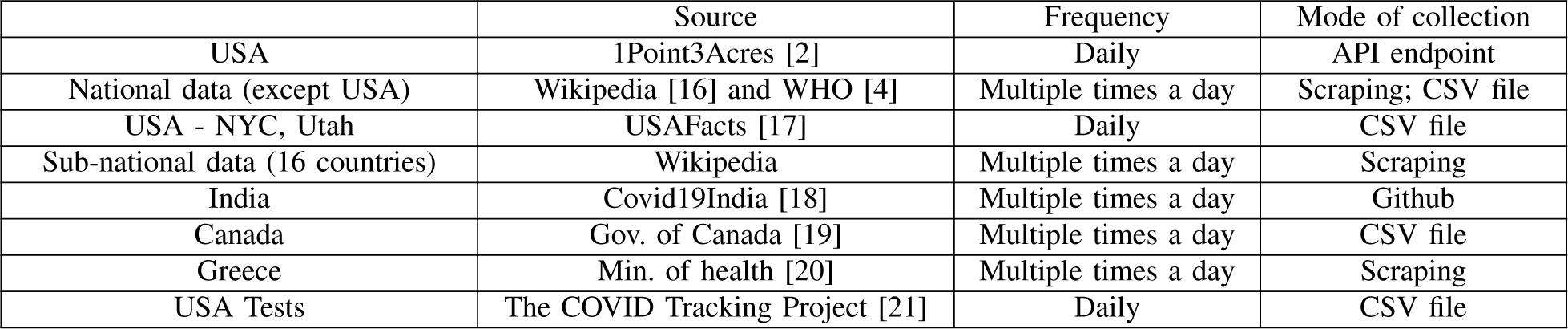
Collection Details of Epidemic Surveillance Data.

#### Demographic data

We use different sources for population data, including Worldometer for country-level population estimates [22], World Population Review for USA state-level population estimates [23], WorldAtlas for China province-level populations [24], Wikipedia for other state/province-level population counts, and Esri Demographics for USA county-level population estimates.

#### GIS data

Polygons for the USA counties are provided by Esri Demographics. Source data for other polygons, e.g., all countries and state/province-level administrative regions, are provided by ADCi [25]. We host these polygons as feature layers on ArcGIS Online [26].

## V. Back-end Architecture

Fig. 3a shows the overall architecture of our dashboard. We present details on the back end in this section, and the User Interface (UI) in Section VI.

As described in Section IV, the data available on our dashboard is multidimensional. At a high level, it includes the surveillance data, demographic data, and GIS data. A design decision made at the beginning of this project was to separate storage of surveillance data from other demographic and GIS data. In particular, the surveillance data is stored locally on our web server and is organized in a spatio-temporal hierarchy, while ArcGIS Online [26] is used to store and access demographic and GIS data for the administrative regions.

### A. ArcGIS Online

ArcGIS Online serves as the GIS server that hosts the feature layers needed by our dashboard. The feature layer is a way to organize collections of separate geographic objects, e.g., administrative regions, buildings, roads, etc., available to the web using ArcGIS. In our dashboard, for easy accessibility, we are using three feature layers corresponding to each spatial level, i.e., one for the world map shown by default on the dashboard, one for states and provinces, and one for USA counties; the feature layers include information such as unique identifier, name and population. Our application fetches data from both the web server and ArcGIS Online, performs a join across datasets on the fly, and uses the joined data for the final visualizations.

Separating constantly evolving surveillance data and relatively static demographic and GIS data in a GIS application is an efficient approach and allows us to support a large amount of data. There are several other advantages: (*i*) by keeping the map services on ArcGIS Online relatively static, we avoid the need to update feature layers, minimizing service outages. (*ii*) The map data only needs to be fetched once, reducing the data transfer between the application and end user to a small amount of data for each new request. This reduces the load on ArcGIS Online, and makes our application scalable for supporting simultaneous requests.

### B. Surveillance Data Pipeline

The surveillance data is collated, processed, and augmented with a robust data pipeline which helps to create an all-in-one comprehensive data hub for all temporal and spatial resolutions. We organized the entire data pipeline in a three-layered approach to effectively achieve the 6Cs as described in the previous sections (See Fig. 2 and Fig. 3b). They are:

#### Layer 1: Curation Layer

This layer focuses on the Current, Curated, and, partially, the Computer-readable elements of the 6Cs standard. We deployed an automated approach for pulling the data from the different data sources every hour and storing it on our clusters, adding a corresponding timestamp (in UTC) for each entry. This helps us to maintain the historical snapshots, and allows users to have access to data from any time period. This also helps ensure that the displayed data is always the latest. The scraping of data from unstructured sources, like webpages and reports, acts as the first step towards making the data Computer-readable.

#### Layer 2: Core Layer

The Core layer is an integral part of our workflow where a huge amount of processing, validation and correction takes place. The diverse sets of raw data stored on our clusters is first combined into a standard and consistent format in accordance with the required spatial and temporal resolutions. This includes a hierarchy with three levels of files each for global data, USA county data, and admin1 regions data. In each of these branches, the data is further organized according to temporal variability, where each file corresponds to the data for a specific day.

Furthermore, data generated by the Core Layer is mapped with FIPS/ID in order to standardize and correctly identify the location. This is specifically essential for the county and admin1 levels where different regions might have a subregion with the same name. Data obtained from multiple sources is very noisy because each data source follows their own data format standard. We have manually identified and mapped each region name with its corresponding ISO3 standard, and also encoded the region names into UTF-8, inspired by the suggestions presented in Addressing the EpiData Challenges [27]. This is a challenging step, since most regions have different languages and encoding, especially from the official sources as they are intended for the local population; it involves a significant amount of effort to manually detect when a new admin1 region is added to our data corpus. With this step, we have completely achieved Consistency of the data in terms of format and historical data. With the standardized Name and ID, the loop of Computer-readability is closed.

The next step is to make sure the data mined from the trusted sources is indeed correct. We do several sanity checks and validations of our data, including checks for a wide range of edge cases and areas where an error might be possible. We then manually correct the data for the identified alerts and warnings to the maximum extent possible by verifying the potential source of the error. A log is maintained for the entries where a resolution was not reachable, thereby achieving our standard for Correctness. This processed, validated, standardized and corrected data is then moved into a central database which serves as our internal modeling and analysis dataset.

#### Layer 3: Augmented Layer

We then augment the data by adding several other derived metrics that help present the overall picture of the pandemic. The augmented data is reflected in the dashboard, and includes the calculation of active cases, the daily change for all the metrics, calculating estimated recovered and estimated active cases, and population-based “per 100K” numbers for all of the metrics. All along our data pipeline, we make sure we properly differentiate between unknown and zero values. This ends the data pipeline, and the prepared data is readily available to be loaded into the front-end for visualization and analysis, thereby completing the loop and achieving Comprehensiveness. The data is populated to our production dashboard multiple times per day in order to present visualizations that are as current as possible.

### C. A Day with the Dashboard : the Data Update Workflow

As shown in Fig. 3b, the data pipeline is a combination of automated data curation procedures that run at a fixed interval, along with some manual steps that are performed each time the data is loaded to production. The data update to the front-end is done multiple times a day, typically starting at 7 AM and extending until 9 PM Eastern time, with a workflow that repeats every 3 to 5 hours (See Fig. 4). During one time step of the update workflow, the logs at the curation layer are checked for potential issues. Since this is an emerging infectious disease, the format of upstream data sources may not be consistent and there could also be an outage at one of the data sources. All of these issues are automatically logged by the data pipeline, and if any issues are detected, we update the scripts used for automated curation of the data accordingly.

Furthermore, validation and sanity checks are executed which issue warnings for scenarios like: a decrease in confirmed cases, deaths, or recovered counts; a significant increase in cases which are above a certain threshold; the appearance of undocumented regions that may have no FIPS/ID and duplicate entries; and stale data for a certain time period, etc. This is a challenging and time-consuming step, especially when there is an official revision from the health department or administration of a region, as this requires data updates to be performed retroactively. After investigating, fixing, and documenting these potential issues, the data update is pushed to a development site. It is again visually inspected, with some exploration, comparison of total counts with other official sources, and a few other test cases.

This human-in-the-loop visual inspection has proven to be an important step, since the sanity checks occasionally fail to identify some issues, and it is an evolving process to make it robust as we go. It also helps to identify those issues that cannot be validated automatically with the data available to us. As a recent example, the United Kingdom’s COVID-19 cumulative death toll was revised downward on August 13th, 2020, by more than 5000. In this particular case, our data sources did not pick up the downward revision, and this made the total deaths on our dashboard seem much higher than on other COVID-19 dashboards. A human-in-the-loop investigation helped to identify the issue so we could fix it before pushing the data to production. This set of steps are repeated for every update on each day.

## VI. Front-end UI design

The User Interface is an important component that complements all the efforts put into the data by making it accessible to a larger audience. A sophisticated data source which is not easily accessible to its non-technical users would limit its potential. True Comprehensiveness can only be achieved when the vast information present in the data is well-conveyed. Realizing the importance of the UI, we have designed it with the goal of providing a one-stop solution that is easy to use and which has rich visualizations accessible to a variety of users with diverse preferences.

To cater to a variety of user needs, we developed a three-way approach for the spatio-temporal exploration of the data i.e., through a choropleth map, charts, and a data table. The map and charts also provide rich visualizations. The Curated spatial data, which is available at multiple resolutions, is presented using a hierarchical approach that follows the principle of “Overview first, zoom and filter, then details-on-demand” introduced by Ben Shneiderman [28]. This means that, initially, the overall picture of the pandemic across the world is displayed, and from there the user can navigate back and forth from one spatial level to another via multiple paths. Taking all this into consideration, the UI is divided into three panels: Control Panel (Header), Information Panel (Charts, Data and Analytics) and Map Panel.

### A. Dashboard Panels

#### Control Panel

The Control Panel (Header) provides temporal exploration functionality through a movie-style time slider or a date selector, spatial exploration through a dropdown list of the countries, and the option to render the choropleth map with different attributes such as Confirmed, Deaths, Estimated Recovered and Estimated Active. It also provides links to more options and toggles to switch between Cases/Testing, Total counts/Per 100K counts, and World map/County map views.

#### Information Panel

The Information Panel, as seen in Fig. 5a, provides the charts and summary for the selected region. The interactive charts include cumulative and incidence numbers with an option to turn an attribute on or off, i.e., Confirmed, Deaths, Estimated Recovered and Estimated Active. The chart can be zoomed in/out, and detailed information is presented in a tooltip window when a user hovers the mouse pointer over a data point on the plot. The interactive data table, which has all the fields of the Augmented layer in the data pipeline, has a SQL-like query tool which allows users to focus on regions of interest, and a ‘Filter’ text box to allow them to limit results to records with the selected region name. The region names in the data table are clickable, which will take the user to the next spatial level if supported, i.e., from the national level to the state level, and then to the county level. The advanced analytics, as discussed in Section VII, gives the users the ability to ask questions related to the pandemic and get answers through data and charts.

#### Map Panel

Our dashboard’s landing page shows a world map at the country level, with the exception that USA and China are shown at the state/province level; we also have additional display layers to support county-level rendering for USA, and state/province level rendering for a total of 20 countries. Unlike most other dashboards that use points to represent the regions, we use actual maps/polygons of administrative regions. The Computer-readability in the data has helped to easily do the geospatial mapping. Each spatial level is rendered with a choropleth map using its Estimated Active count by default, and the map can be zoomed in/out. For a selected region, a pop-up window is provided to show its data, along with a navigation option to change the display level of that region.

One unique feature is the multiple ways a user can select a desired region. Spatial exploration can be done through the dropdown list selector, the SQL-like query tool, the filter text box, navigating through the data table, as a question in the analytics, and selection on the map.

### B. Implementation Details

Our dashboard is a Single Page Application which loads data and default map rendering into an HTML page for the Current data; it is dynamically updated whenever the user interacts with the application. On the application level, our dashboard leverages existing development APIs to provide a basemap layer, and incorporate interactive charts and the data table within a system that is specifically designed to be loosely coupled.

#### 1) ArcGIS API

Unlike most other dashboards that are built using the configurable ArcGIS Dashboards template, the front end of our dashboard is primarily built on top of the ArcGIS API for JavaScript [29]. Although this was a more challenging approach at the beginning of the project, once we completed that implementation and built the framework, it gave us more flexibility for creating a customizable and functionally rich application. For example, we could add a movie-style time slider to effectively show spatial variation over time, it allowed us to use amCharts 4 [30] for advanced data visualization, and we are able to use DataTables for jQuery [31] to allow users to explore the data in a tabular form. This is the reason why our dashboard looks quite different from most of the other dashboards, and why it has such an extensive set of unique features.

#### 2) Responsive Web Design

Our dashboard allows users to access the application on any handheld device, such as an iPad, Tablet, or Mobile phone. To tackle this challenge to make the application available on a variety of screen resolutions, we used the Responsive Web Design approach to design and develop our dashboard. This allows the front end to respond to the user’s behaviour and environment based on their screen size, platform, and orientation. The practice consists of a mixture of JavaScript, jQuery, Bootstrap, flexible grids and layouts, images, and an intelligent use of CSS media queries.

#### 3) amCharts

amCharts 4 is a go-to library for advanced data visualization which provides a simple, yet powerful and flexible, drop-in data visualization solution. It includes all basic and advanced chart types, and also supports responsive web design. In particular, we are using line charts and stacked bar charts to visualize incidence and cumulative data, and the comparison plots for the Top K highly affected countries where K can be 5, 10, 15 and 20.(Fig. 5b)

## VII. Analytics

The plethora of information present in the data cannot be effectively and completely conveyed on a single web page without cluttering it. Providing users with the ability to quickly and directly get answers to generally asked questions will further minimize the need for them to understand the epidemic terminology, allowing them to spend more time navigating the application. This is how ultimate Comprehensiveness can be achieved, and it also helps to serve the diverse information needs of the public. In order to achieve this, we support interactive queries for analytics, where a user can ask a question in plain conversational English, which the system attempts to interpret, then answers directly with a plot and data as appropriate (See Fig. 5c).

### A. Methodology for Analytics

We have identified that a question related to an epidemic will typically be composed of 4W1H (which, what, where, when, how) as shown in Table II. This provides a basis for processing the data and responding effectively to user queries.

**TABLE II:** 4W1H Structure of Epidemic Questions.

| Which | What | Where | When | How |
| --- | --- | --- | --- | --- |
| Confirmed<br>Deaths<br>Est.Recovered<br>Est.Active<br>Recovered<br>Active | count<br>greater than / less than<br>top N / bottom N<br>highest / lowest | India<br>Virginia<br>Queens<br>United States<br>Lombardy<br>World | March 15<br>Today<br>July 10 | Cumulative<br>Incidence |

Some examples of the questions supported by the tool include: “Cumulative deaths count in India on July 10”, “Cumulative confirmed greater than 100000 in United States today”, “Top 5 incidence deaths in Virginia state on April 15”. The 5W1H acronym is quite popular in research, journalism, investigations etc, and ‘Who’ replaced with a more relevant ‘Which’ for epidemic data. The other W that is not supported in our 4W1H formulation is ‘Why’. This category of question is not trivial to answer from the surveillance data without the context of the real-world situation, which is often very complex. The default values for each of the 4W1H, if the user doesn’t provide them explicitly, are taken as follows: Which - Confirmed, What - count, Where - World, When - Today, How - Cumulative. The system implementation of 4W1H can search the words in the question for each of the 4W1H keywords in the search space. The search space include 6 possibilities for ‘Which’, 7 for ‘What’, around 3750 regions (210 countries, 350 states, 3200 counties) for ‘Where’, the no.of days since pandemic start for ‘When’ and 2 for ‘How’.

### B. Semantic Textual Similarity and The User Query Dataset

To allow users to freely ask questions without restricting them to certain words as shown in Table II, a state-of-the-art model like SentEval [32] can be used to compute semantic similarity of the tokens in the question against the words within the search space, and to return the result for the match which had the highest similarity score. Whenever a question fails to get a result directly from the search space, this can be invoked and the relevant results can be displayed if the mean similarity score meets an empirically determined minimum threshold. If it fails to meet the minimum threshold, it will be considered a failed query and the user can be alerted that the question could not be answered.

To keep track of the user questions and the status of the results, we save them into a database along with other details like ‘Date’, ‘Time’ and ‘Result (Pass/Fail)’ for each question. This helps us identify which queries are failing, allows us to broaden the scope of our methodology if required, and also guides improvement to the sentence similarity model. Furthermore, there is a feedback option where the users can submit a response if they are satisfied with the results of their question which we can also use to improve the system. This will be a first-of=its-kind dataset containing real-time epidemic questions from users located all over the world. This data could potentially open up new areas of research on understanding the data needs of the public, allowing applications to more closely target their responses during a pandemic.

### C. Current Implementation and Future Work

Currently, we support spatial questions for the current state of the pandemic, i.e., 3W1H with the exception of ‘When’. Whenever a question cannot be resolved within the given search space, the system catches the exception and provides the user with a warning that their question could not be answered. We are expanding the supported input space by manually reviewing the failed queries from the User Query dataset and incorporating changes where feasible. This includes support for sets of synonyms, such as: [greater than, more than, higher than], and [United States, US, USA, America], etc.

We are in the process of adding support for the temporal ‘When’ questions, and exploring state-of-the-art sentence similarity models to incorporate them in future releases. We are also exploring the expansion of 4W1H input space to add support for Tests to the ‘Which’ handling, and for Peak, 7-day Moving Average, Test positivity rates, etc. to the ‘What’.

## VIII. Timeline

Our dashboard was not built in a single release; we went through numerous design iterations and feature improvements to achieve the 6Cs and the other goals. The data sources we depend on have evolved over time as well, requiring us to find new sources as new features were added. Development of this application continues to be an evolving process, and the timeline for our project thus far can be divided into four significant phases as described in Fig. 6.

**Fig. 6:**
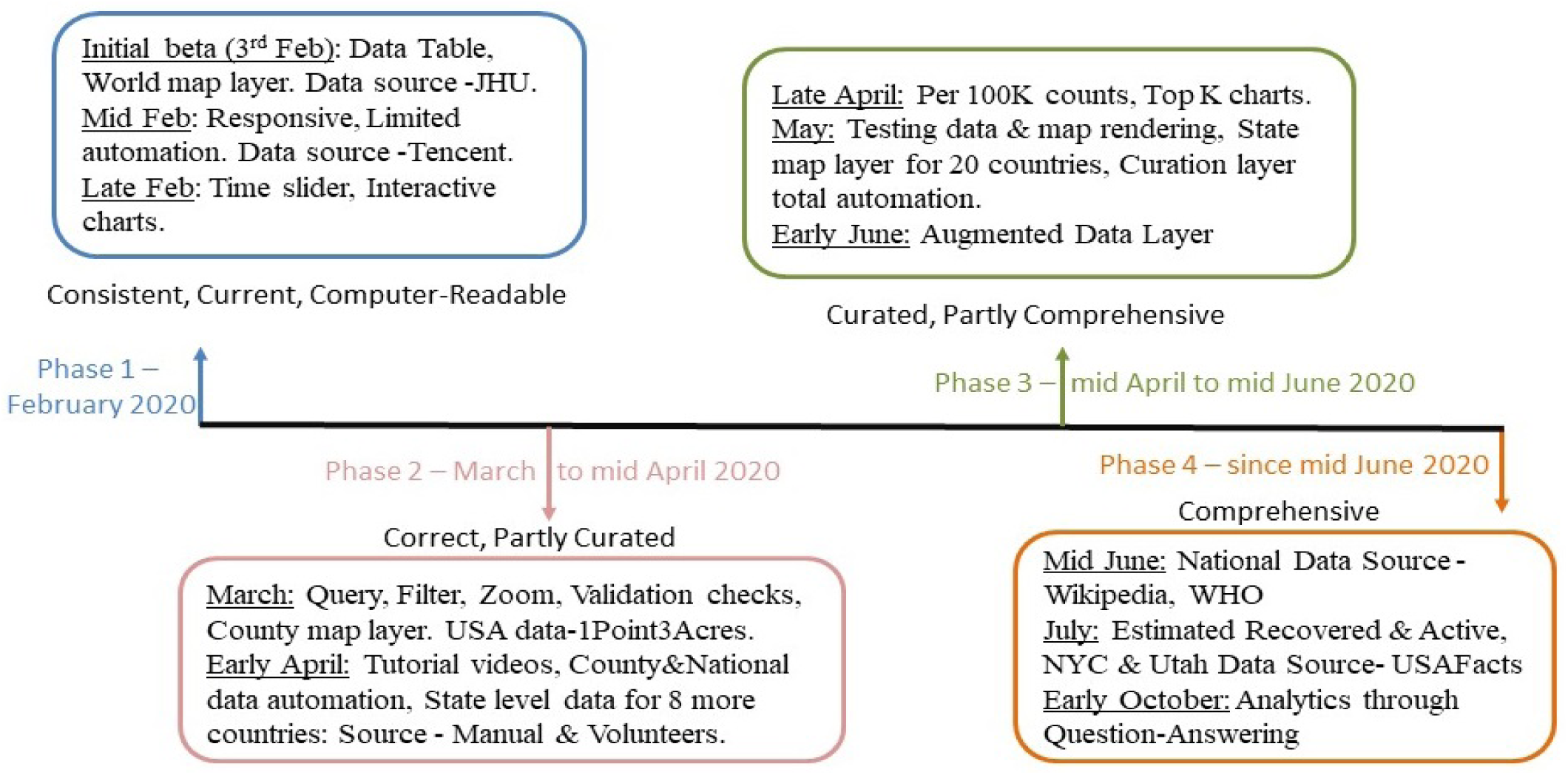
Project Timeline. The course of the development of the project can be broadly divided into four phases; this timeline details which features were added at each phase, and when each of the Cs were achieved.

### A. First Phase (February 2020)

The initial beta release of our dashboard was deployed on February 3, 2020, with a limited feature set, i.e., only the data table and country-level rendering. USA and China were the only countries which had state/province-level data and geospatial visualizations. The website was not mobile-friendly at this point; however, as we analyzed usage information, we noticed that more than 50% of the users were using mobile devices. This motivated us to make development of a more responsive and mobile-friendly interface our top priority; this release went live in mid-February. Our initial data source was JHU [1], and data curation was a manual process at that time. Starting in mid-February, we switched our primary data source to Tencent [33] as it’s more consistent, and implemented a limited automated workflow for the data curation and integration; while we continued to use a manual process for curating the USA, it required a lot of repeated steps to keep the data current. By late February, the time slider and interactive charts had been added, and we ensured that the data was Consistent, Correct, Current and Computer-readable.

### B. Second Phase (March to mid April)

As the COVID-19 pandemic started to unfold, we focused primarily on three areas: more data coverage, new features, and extensive automation of the data curation process. Starting from early March, we were one of the first dashboards to show USA data at the county level using data from 1point3acres [2] as the source. Querying and filtering the data by region and enabling zoom in/out functionality on multiple spatial levels helped users to explore the pandemic situation at a granular level, e.g., checking the situation at their loved ones’ locales. To allow our users to get the most out of our dashboard, we created a series of tutorial videos explaining the features and how to explore and interpret the data. During this phase, we transitioned to complete automation of the Curation layer, and we added various validation steps that were run before curated data could be moved to the core layer. To ensure data quality, we performed a manual review before each data update. We were also fortunate to have volunteer support for data collection at the state/province level in some countries, including Germany, Chile, Brazil, Colombia, Greece and Argentina – a step forward towards the ‘Curated’ goal.

### C. Third Phase (mid April to mid June)

We continued to improve our dashboard with more data and features. We added the data and visualization for COVID-19 Testing, added per 100K counts, added the Top K country charts, and many minor updates. We automated the state/province-level data curation, and extended the list of countries with state/province support to 20, with map rendering through a new third map layer; these countries included Argentina, Austria, Brazil, Belgium, Canada, Chile, China, Colombia, Germany, Greece, India, Italy, Mexico, Peru, Portugal, Saudi Arabia, South Korea, Sweden, Switzerland and USA. We also extended the data pipeline with the augmented data layer to ensure the consistency of core layer data while adding more derived metrics for the application. At this stage, we achieved the Curated and Comprehensive goals, thereby achieving all of the 6Cs.

### D. Current Phase (since mid June)

In the first half of June, we switched the data source for National data to Wikipedia and WHO. As mentioned in IV-A, we began providing Estimated Recovered and Estimated Active numbers in addition to the much less accurate reported recovered and active counts. We believe this will provide a better picture of the current situation, and will be helpful for decision-making at the individual level. Finally, we added Advanced Analytics, which allows users to ask questions regarding the pandemic using natural language, and provides the answers through data and plots; we believe this is a great step towards making the dashboard more Comprehensive.

## IX. Utility of the Dashboard

### A. Data storytelling

An important application of our dashboard is in support of *data storytelling*. Data storytelling is the art of developing a narrative based on a data set, incorporating visualizations and analysis tools so viewers can make solid, well-supported interpretations; it is quite popular in the fields of data science [34] and data journalism. The Analytics of the dashboard, along with its historical data and interactive visualizations, make it easy to gain insights that facilitate data storytelling. An excellent illustration of this concept is a blog post by Tomas Pueyo, who has produced an extremely interesting narrative on the COVID-19 pandemic [35]. We provide here a simple and brief illustrative example of how someone could compose a data-driven narrative on the rise of COVID-19 cases in the USA and Europe using our dashboard.

#### Contrasting the pandemic trajectory in the US and Europe until August

Although COVID-19 was first reported in Wuhan, China, Europe was also an early player in the pandemic; the first positive case in Europe was reported in France, and it appears the patient was infected in mid-December. The disease then quickly spread across Europe, and three of the four largest countries, Italy, Spain, and France, showed significant rises in cases starting in the first week of March. In contrast, with the exception of a small cluster of cases in January, the pandemic began rather late in the USA; however, by the second week of March, things were picking up. Figure. 7 shows the curves of daily confirmed cases in USA and Europe. Italy and Spain were among the top 5 countries in the world by March 12. National lockdowns were implemented in many of the European countries – Italy on March 9, and a state of emergency was declared in Spain on March 13 – and the effects of these lockdowns were noticeable by April 5. By this time, Europe had started to see a turnaround, and had largely controlled the pandemic by mid-May; even by that time, there were a significant number of deaths and confirmed cases. Back in the USA, the initial epicenter was in Seattle, Washington, but soon moved to New York state. By April 15, there were around 13,000 total deaths in New York, with approximately 10,000 confirmed cases each day. Initially, the cases were restricted to a shortlist of states, like New York, New Jersey, California and Michigan. By the end of April, however, New York began to regain control over the pandemic, with incidence cases dropping to 4500. By Memorial Day, one could see the daily case count was substantially lower, hovering around 1500 cases. Then came Memorial Day: things were under control, but restrictions started to loosen, and this soon caused a resurgence. By this time, the pandemic was widespread, and many states started to see an uptick in cases. While New York seemed to be flattening the curve, states like Florida, Texas and California had the highest case counts. The nationwide daily case count quickly rose from 20,000 cases per day to 45,000 at the end of June. It further rose to a daily case count of 70,000 cases by mid-July, with the above-mentioned three states each recording over 10,000 cases per day. At this time, the daily case count in Europe was around 11,000, and New York had also kept the pandemic under control, seeing less than 1000 cases a day. But Florida, Texas, and California saw significantly lower death rates even as the number of cases approached that of New York (See Fig. 8). To respond to this surge in cases, the states halted further reopening plans, and by early August the pandemic epicurve seemed to be on a downtrend.

**Fig. 7:**
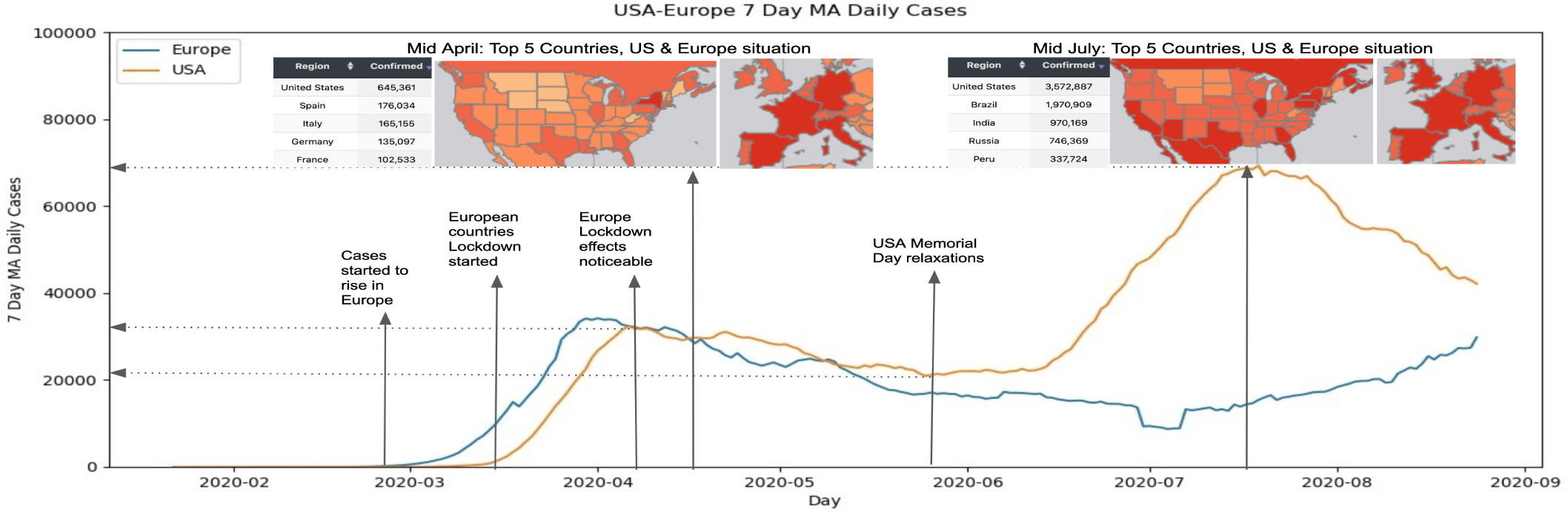
Case counts rose (and fell) in the USA and Europe in response to significant events over the course of the pandemic. The snapshots illustrate the spatial impact of the pandemic in different geographical regions during times of peak activity

**Fig. 8:**
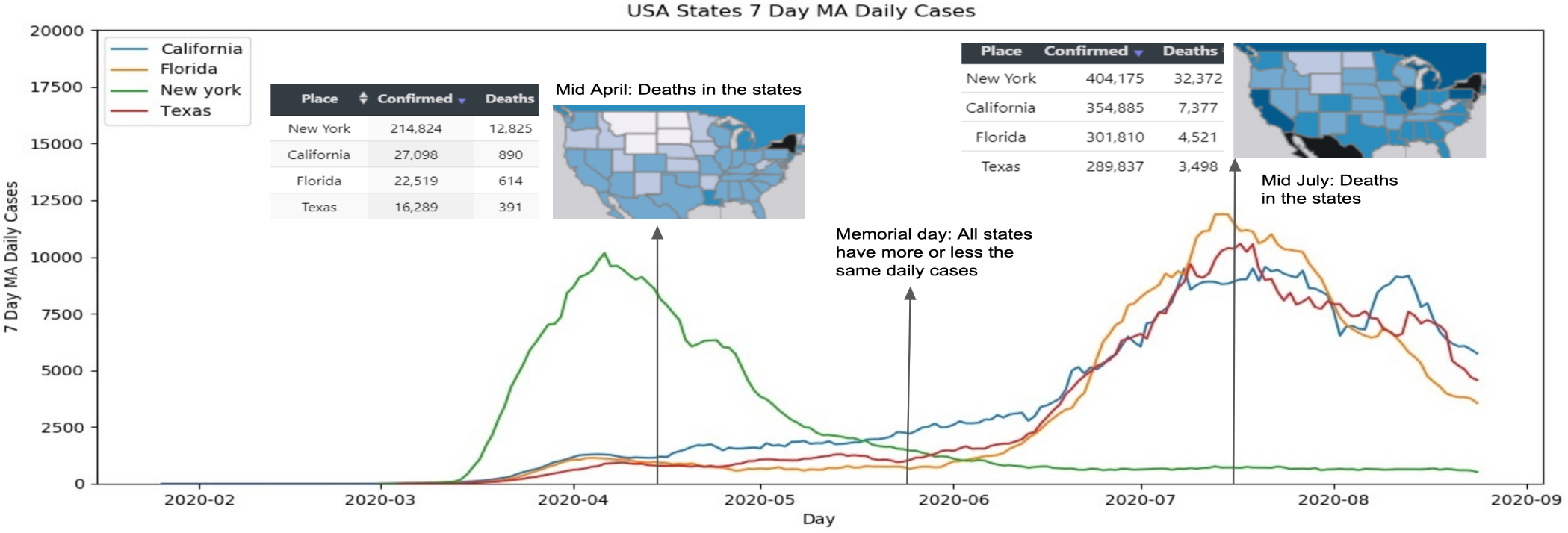
Data storytelling: USA States Timeline for 7-Day Moving Average of Daily Cases for New York, California, Florida and Texas. The snapshots of the choropleth maps show deaths in Mid-April and Mid-July as compared with the cases.

### B. Extensive use by various organizations

The application and the back-end data have been used by a large number of analysts, researchers, and laypeople. Our group uses the data to support federal agencies (DoD, CDC), our state (Virginia) and local public authorities (local health districts and our university) as they respond to the pandemic. The data is also used to drive our predictive models, which we use to produce counter-factual analysis, and answer policy questions, including contact tracing, resource allocation and augmentation, and campus reopening and management. See [36] for reports that the Virginia Department of Health (VDH) releases based on our work.

Several other groups use our dashboard and associated data as well. We list just a few to illustrate its broad use: (*i*) it is listed as a part of the NIH MIDAS data catalogue [37], the ESRI COVID-19 GIS Hub [38] and the 2021 Coalition for Academic Scientific Computation (CASC) brochure; (*ii*) it is used by several groups at DoD; (*iii*) it is used by local authorities, including in Bay County [39] and Panama City Beach [40] in Florida, where our active case counts are used as one of their thresholds for allowing vacation rental reservations.

### C. Web Traffic

During the initial phase, we had around 40,000 users in total, and the top 3 countries for web traffic were the United States, Germany and China. By the end of the second phase, we reached over 750,000 unique users, and the top 3 countries for web traffic were the United States, Canada and Germany. By late October, there were over 1.13 million unique users. The top 3 countries that made up the largest portion of users are currently the United States, India and Canada. Overall, two-thirds of the users are from the United States, and the average time spent on our dashboard is around two minutes. The period of maximum engagement was the first week of April, with a peak of 80,000 views and 50,000 unique users on a single day. The overall returning user percentage stands at 19% and it was 11%, 19%, 31% and 22% respectively during our four phases as described in the timeline.

## X. Discussion and Conclusion

### A. Challenges

Getting data from reliable sources and conforming to the 6Cs is the most challenging task in building and maintaining our dashboard, yet worth the effort to maintain the quality. Changes in the Terms of Use at some of our upstream data sources was another big hurdle to overcome, and we had to switch data sources at times as a result. We faced other challenges, especially with the state/province-level data, which lacks a standard format and computer-readability in some of the upstream sources. The difference in the naming of regions also makes it difficult to identify corresponding spatial coordinates, and required a manual effort to map the names. The amount of work involved has limited our ability to expand state-level coverage to additional countries.

With Recovered numbers not being reported consistently across all countries, it has been hard to rely on reported Active case counts, which should be the ideal metric for understanding the current situation. This led us to develop an algorithm for computing estimated recovered values based on available research and studies. Inconsistencies in the definitions of “case” and “death” across the regions also created issues, with some regions reporting presumed case/death counts, while some data sources report only confirmed cases/deaths. This also led to retrospective data corrections in some cases. There have been instances where cases were defined under unknown regions, as well as cases where more than one region was clubbed together. These have imposed some challenges, especially when the data is intended to be used in modeling; these situations had to be handled on a case-by-case basis.

With the rapidly evolving pandemic and short release cycles, robust testing of our application also turned out to be a challenging task. Building our dashboard on top of the ArcGIS API for JavaScript was a key decision we made at the beginning of the project. This made our dashboard stand out in terms of how it looks and the unique features we are able to deliver. This demanded a significant effort at the beginning to build the framework.

### B. Lessons

The onset of COVID-19 led to the creation of many special-purpose data collection and visualization efforts that focused on specific aspects of the pandemic, such as current case counts or the status in a particular region. We see the need for a more general-purpose, all-in-one central repository that can provide broader coverage of the pandemic at a granular level of spatio-temporal detail; corresponding data update access could be granted to public health officials, journalists, and researchers to make this a global community-response resource promoting timely decision-making and research.

COVID-19 has shown the importance of easy-to-use and interactive visualizations of the epidemic data to help the public stay informed. We learned that, during an emerging infectious disease, speedy response is essential, and a ready-to-deploy dashboard without much of a development effort could save critical time. Our current design of the application architecture, guided by the 6Cs of epidemic data management, is flexible and adaptable and could be deployed quickly in case the need arises for another specialized pandemic site in the future.

We also provided a support email for feedback, suggestions, and questions from the users. It turned out to be very useful for understanding public opinion, and over time we performed many rollbacks, upgrades and changes based on the active feedback received. The tutorial videos attracted many viewers and were widely appreciated.

### C. Limitations

Due to the challenges in the collection and maintenance of the state/province-level data, with the inconsistent formats at upstream sources, our dashboard currently only supports 20 countries at this level of granularity. Also, while a large number of countries do not have this data reported, others provide it in the form of text-based reports or press releases, hence making it difficult to extract on a continual basis; this leaves room for improvement for Curation.

### D. Conclusion

This paper presents our epidemic surveillance dashboard in support of the COVID-19 pandemic planning and response. We propose the 6Cs metrics as a possible standard for epidemic data management. Our application has been used extensively by users globally. The need for such systems is clear, and we hope it spurs further discussion on how data can be collected and shared during such pandemics.

## Data Availability

The COVID-19 Surveillance data is openly accessible for viewing at our Dashboard

https://nssac.bii.virginia.edu/covid-19/dashboard/

## Acknowledgements

The authors would like to thank members of the Biocomplexity Institute and Initiative; Paula Stretz and Richard Beckman for useful discussion, suggestions and testing in the development process; Drew MacQueen, from the Scholars’ Lab at UVA Library, for helping with the feature layer (map) used in the dashboard; and members of the team at Persistent Systems, for their assistance in the user interface design and testing. This work was partially supported by the National Institutes of Health (NIH) Grant 1R01GM109718, NSF BIG DATA Grant IIS-1633028, NSF DIBBS Grant ACI-1443054, DTRA subcontract/ARA S-D00189-15-TO-01-UVA, NSF Grant No. OAC-1916805, US Centers for Disease Control and Prevention 75D30119C05935, and a collaborative seed grant from the UVA Global Infectious Disease Institute.

## Footnotes

1 This paper will appear in 2020 IEEE International Conference on Big Data, December 10–13, 2020, Virtual Event, USA

